# MASSIVE SCABIES OUTBREAK IN ROHINGYA REFUGEE CAMPS, COX’S BAZAR: EPIDEMIOLOGY AND IMPACT OF A MASS DRUG ADMINISTRATION (MDA) CAMPAIGN – A RETROSPECTIVE STUDY

**DOI:** 10.64898/2025.12.05.25341689

**Authors:** Charls Erik Halder, Md Abeed Hasan, James Charles Okello, David Otieno, Emmanuel Roba Soma, Partha Pratim Das, Md Mostafizur Rahman, Md Atiquzzaman, Hamim Tassdik, Dickson Wafula Barasa, Julekha Tabassum Poly, Abdullah Al Mamun, Abu Toha Md. Rizuanul Haque Bhuiyan, U Maung Prue

**Affiliations:** Migration Health Division, International Organization for Bangladesh (IOM), Bangladesh; Epidemiology Team, World Health Organization (WHO), Bangladesh; Refugee Refugee Relief and Repatriation Commissioner (RRRC), Bangladesh

**Keywords:** Scabies, Epidemiology, Mass Drug Administration, MDA, Rohingya, Disease Outbreaks, Communicable Disease Control, Bangladesh, Cox’s Bazar

## Abstract

**Background:** Since 2021, Rohingya refugee camps in Cox’s Bazar, Bangladesh, have experienced a significant and concerning increase in the prevalence of scabies. In response to the massive outbreak, an extensive Mass Drug Administration (MDA) campaign was conducted from November 29, 2023, to February 01, 2024.

**Objectives:** The objectives of the study were: a) to determine the epidemiological characteristics of the scabies outbreak in the Rohingya refugee camps, including magnitude, age-sex distribution, and attack rate; and b) to evaluate the impact and durability of MDA in reducing the burden of scabies in the refugee camps.

**Methodology:** This was a retrospective observational study that utilized deidentified and anonymized data from 35 health facilities of the International Organization for Migration (IOM) spanning the period from January 1, 2021, to December 31, 2024.

**Results:** A total of 384,852 cases of scabies were reported, with an overall attack rate of 5,562.59 scabies cases per 10,000 population over 4 years. Females had a slightly higher case proportion (53.51%) and attack rate (ARR: 1.104, p < 0.001). Children under 5 years (36.31%) had the highest burden and attack rate, about twice the overall attack rate. Using this age group as reference, the attack rate declined significantly with increasing age, 5,561.85 among adolescents (ARR: 0.496, p < 0.001) to 4,377.88 among individuals over 60 years (ARR: 0.390, p < 0.001). 77% of the cases were reported among the Rohingya refugees; however, the attack rate was higher among host communities (7,721.34 per 10,000 vs. 5,138.24 per 10,000 among refugees). Overall, 5.80% of scabies cases were associated with secondary bacterial infections. Interrupted time series (ITS) analysis showed a sharp decline in scabies cases (from 1,885 cases per week to 0, *p* < 0.001) following the initiation of MDA. The decline caused by MDA persisted for 6 months, after which an upward trend was observed (with an increase of 72.04 cases/ per week).

**Conclusion:** The study revealed an extremely high burden of scabies among the Rohingya refugees and the adjacent host community. MDA was an effective approach for a rapid and substantial reduction of the disease burden. However, the impact cannot be sustained unless the underlying factors of the scabies outbreak are addressed.

## INTRODUCTION

Scabies, classified among neglected tropical diseases, impacts over 400 million individuals annually (WHO) (1,2). It is a parasitic skin disease caused by the Sarcoptes scabiei mite, which intricately burrows into the skin, lays eggs, and incites severe pruritus (3,4). Scabies is primarily transmitted through direct skin-to-skin contact. Secondary bacterial skin infections from scabies are a significant risk factor for immune-mediated complications, including acute post-streptococcal glomerulonephritis (manifesting as kidney disease), rheumatic heart disease, and death (2–6). While scabies is endemic globally, its prevalence is notably higher in hot and tropical countries as well as in regions characterized by high population density (2). Consequently, refugees and displaced communities confront an elevated risk of transmission due to overcrowded living conditions, insufficient access to water, sanitation, and hygiene facilities, challenges in laundry of clothes and bedding, limited access to healthcare, shortage of scabicidal drugs, and socio-cultural factors such as language barriers and stigma (7,8). The prevalence is heightened in Rohingya refugee camps due to overcrowding and limited access to essential healthcare. A high burden of scabies has been reported in other refugee settings across the globe, such as Greece (13,118 reported cases over four years) and Duhok, Iraq (up to 20% prevalence), highlighting the global burden of the disease in similar humanitarian contexts (9,10). Following decades of persecution and repeated displacement, approximately 939,344 Rohingya refugees fled to Bangladesh in August 2017. As of December 31, 2023, they reside in densely populated camps in Cox’s Bazar, where overcrowding, fragile shelters, poor WASH (water, sanitation, and hygiene) infrastructure, and harsh monsoon conditions significantly increase vulnerability to infectious disease outbreaks, including scabies (11). The camps have experienced multiple outbreaks or surges of infectious diseases, including diphtheria, measles, COVID-19, and Acute Watery Diarrhea (AWD)/cholera (12–16). Since 2021, the refugee camps have witnessed a significant and concerning surge in the prevalence of scabies. The high burden of scabies has led to an increased demand for heightened medical attention and additional resources, exacerbating financial strain to an already strained health system within this protracted humanitarian crisis. Simultaneously, it contributes to disruptions in daily life, potentially hindering social interactions and economic activities within the refugee community.

Despite scabies being a critical concern in refugee and displacement settings due to its negative impact including disability, stigma, and exacerbation of poverty globally, there are only a few studies available that have documented the epidemiology and management of scabies in such settings (8,10,17,18). Understanding the epidemiology of scabies, including its patterns and magnitudes of outbreaks, is essential for effective public health planning, prevention, and response.

In response to the massive outbreak of scabies, from November 29, 2023, to February 01, 2024, an extensive Mass Drug Administration (MDA) campaign was conducted to targeting nearly 1 million beneficiaries residing in 33 camps in Cox’s Bazar and Bhasan Char Island. This campaign was recognized as the world’s most extensive MDA campaign for scabies treatment and prevention (1,2). MDA is a public health strategy that involves providing treatment to entire population, irrespective of their disease status, and has been practiced extensively in recent decades as part of a global effort to control and eliminate neglected tropical diseases (19). For the MDA campaign in Cox’s Bazar, Ivermectin 3mg tablets and Permethrin 5% cream (for contraindicated cases) were used.

A systematic review and meta-analysis of 11 reports on the impact of MDA found it to be a highly effective strategy in reducing the prevalence of scabies (20). However, the study concluded that further research is warranted to understand the effectiveness of MDA regimens in larger populations and to determine the durability of impact. Therefore, in addition to explaining the epidemiology and magnitude of the outbreak, our study evaluated the impact of the MDA campaign and its durability.

In summary, the objectives of the study were: a) to determine the epidemiological characteristics of the scabies outbreak in the Rohingya refugee camps, including magnitude, age-sex distribution, and attack rate; and b) to evaluate the impact and durability of MDA in reducing the burden of scabies in the refugee camps. The findings of the research will assist health programmes and policymakers in predicting the scale and characteristics of scabies outbreaks, as well as the effectiveness of public health interventions, thereby facilitating proper planning for the effective integration of scabies prevention and management within the essential health service model.

## METHODOLOGY

### Study design

This was a retrospective observational study that used deidentified and anonymized data from the International Organization for Migration’s (IOM) scabies patient database.

### Study site and population

The study was implemented in Rohingya refugee camps in Ukhiya and Teknaf in Cox’s Bazar. The study population included all individuals who were clinically diagnosed with scabies and received care at 35 IOM-supported health facilities in Ukhiya and Teknaf, Bangladesh, between 1st January 2021 and 31st December 2024. Scabies caused a significant outbreak during this period in the refugee camps that prompted a substantial response from IOM. Therefore, these camps were selected for this study to get an in-depth idea of the outbreak.

### Data collection

Deidentified and anonymized aggregated data pertaining to scabies cases were retrieved from the IOM Cox’s Bazar’s health information management record spanning the period from 2021 to 2024. The data included age, sex, camp location, and presence of secondary infection. All data were collected using the IOM Cox’s Bazar OPD form, which was further transmitted electronically using Kobo Toolbox and synchronized at the central database. The data was centrally cleaned and verified daily before being exported to an excel spreadsheet for this study’s analysis.

### Scabies diagnosis and clinical management

All scabies cases were clinically diagnosed at 35 IOM-supported facilities in Ukhiya and Teknaf. MSF clinical guidelines (21) A– diagnosis and treatment manual was used for diagnosis and clinical management of the cases. 5% permethrin cream was mainly used for the treatment of scabies.

### Mass Drug Administration (MDA Campaign)

Between November 29, 2023, and February 01, 2024, WHO Bangladesh, in collaboration with the Government of Bangladesh and Health Sector partners, implemented the world’s largest Ivermectin-based MDA campaign in response to the massive scabies outbreak among the Rohingya refugees in 33 camps across Cox’s Bazar and Bhasan Char (22). According to the WHO, the MDA regimen includes a dose of ivermectin at 200 µg/kg body weight administered biannually 7-14 days apart. The WHO 2023 guidance suggests that ivermectin is not recommended in children less than 15 kg or 90 cm in height, in pregnant women in the first trimester, and with those who have the known allergy to ivermectin or permethrin (23). The MDA consisted of two rounds of doses from November 29, 2023, to February 1, 2024, covering all 33 camps in Teknaf, Ukhiya, and Bhasan Char Island. The campaign provided therapeutics to over 992,500 Rohingya refugees, including 5.2 million Ivermectin tablets and 185,000 tubes of Permethrin cream. IOM contributed 60,000 drugs (Ivermectin, 6 mg) in this campaign, allowing the second dosing of the drug among 33,970 residents in Camp 09 and Camp 20 Extension (22)

### Statistical analysis

Analysis of epidemiological characteristics: The cleaned dataset was summarized into scabies cases and infected scabies by weeks, months, and years through descriptive statistics. Attack rates were calculated annually and overall, using total scabies cases divided by the respective catchment population, expressed per 10,000 person-years. Attack rates were also segregated for years, age, sex, geographical location (camps and Upazila), and type of community (host vs refugee) using available population denominators. UNHCR population data was used as the population denominator. Attack rate ratios (ARR) with corresponding 95% confidence intervals and p-values were calculated to compare scabies burden across sex, age groups, geographic location (Upazila), and community (host vs refugee). P-values were derived by the chi-square test. Proportional morbidity for scabies was calculated as the percentage of all outpatient consultations expressed by month and year.

An Interrupted Time Series (ITS) analysis was performed to assess the impact of MDA on the scabies trend and durability of the impact. The ITS model consisted of pre-MDA, MDA phase, and post-MDA segments to estimate changes in level and trend, from January 2021 to December 2024. Statistical applications, including R, R Studio, and Microsoft Excel, were used for data analysis and visualization, including tabulation, graphical analyses, and regression analyses.

## ETHICAL CONSIDERATION

The research team received ethical clearance (CoxMC/2023/017) from the Ethical Review Board, Cox’s Bazar Medical College Hospital. The development of the research was discussed with, and necessary clearance was obtained from the office of the civil surgeon and the Relief and Repatriation Commissioner (RRRC) health coordinator, who are responsible for overseeing activities within the scope of the refugee crisis in Cox’s Bazar, Bangladesh. This was a retrospective observational study, and thus, only utilized historical anonymized data from the outbreak database. All individual data used in the analysis were anonymized (i.e., no personal identifiers), and only aggregate results were reported. All data presented in the study were gathered during the public health outbreak response; therefore, ethical clearance prior to the data collection was not obtained.

## RESULTS

A total of 384,852 scabies cases were reported from the IOM-supported health facilities in Cox’s Bazar from January 2021 to December 2024. Overall attack rate of scabies over 4 years was 5,562.59 per 10,000, with an average annual attack rate of 1492.82 per 10,000 population (Table 1).

**Table 1:** Distribution of scabies cases and attack rates by demographic characteristics.

| Variable | Category | Scabies<br>Count | Percentage<br>(%) | Population | Attack<br>Rate | Attack<br>Rate<br>Ratio | P value |
| --- | --- | --- | --- | --- | --- | --- | --- |
| Overall | Total | 384,852 | 100.00% | 691,857 | 5,562.59 | 1.000 | - |
|  | Mean age (years) |  | 14.82 ± 16.11 years |  |  |  |  |
|  | Median age (years) |  | 9 |  |  |  |  |
| Sex | Male | 178,917 | 46.49% | 338,672 | 5,282.90 | 1.000 | Ref |
|  | Female | 205,935 | 53.51% | 353,185 | 5,830.80 | 1.104 | <0.001 |
| Age group | 0 – 5 years | 139,747 | 36.31% | 124,634 | 11,212.59 | 1.000 | Ref |
|  | 6 – <18 years | 129,406 | 33.62% | 232,667 | 5,561.85 | 0.496 | <0.001 |
|  | 18 – <30<br>years | 54,485 | 14.16% | 304,642 | 1,788.49 | 0.160 | <0.001 |
|  | 30 – 60 years | 48,118 | 12.50% | 48,002 | 10,024.17 | 0.894 | <0.001 |
|  | 60 years+ | 13,096 | 3.40% | 29,914 | 4,377.88 | 0.390 | <0.001 |
| Upazila | Ukhiya | 283,689 | 73.71% | 530,560 | 5,346.97 | 1.000 | Ref |
|  | Teknaf | 101,163 | 26.29% | 161,297 | 6,271.85 | 1.173 | <0.001 |
| Community | Host | 87,760 | 22.80% | 113,659 | 7,721.34 | 1.000 | Ref |
|  | Refugee | 297,092 | 77.20% | 578,198 | 5,138.24 | 0.665 | <0.001 |

Females accounted for a slightly higher proportion of cases (53.51%) and had a higher attack rate ratio than males (ARR: 1.104, p < 0.001). The highest burden of the cases was reported from children under 5 years (36.31%) and adolescents aged between 6 and 18 years (33.62%). However, the attack rate was significantly higher among the children under the 5-year age group (11,212.59) in comparison to any other age group and more than twice the overall attack rate. Using this age group as reference, the attack rate declined significantly with increasing age, 5,561.85 among adolescents (ARR: 0.496, p < 0.001) to 4,377.88 among individuals over 60 years (ARR: 0.390, p < 0.001).

Geographically, about three-quarters of the cases were reported from Ukhiya (73.71%); however, the attack rate was slightly higher in Teknaf (ARR 1.173, p <0.001). While more than 77% of the cases were reported among the Rohingya refugees, the attack rate is lower than that of the adjacent host communities (AAR 0.665, <0.001).

Figure 1 illustrates a camp-based geo-spatial analysis was performed to determine where scabies cases were located in the refugee camps of the catchment area. Camps are shaded in red based on total scabies case load with blue circles overlaid based on attack rates (per 100,000). Increased circle size equals increased attack rate. The following camps had the highest attack rates: Camp 20 Extension (18,259 per 100,000), Kutupalong RC (18,084) (33,979 cases), Camp 13 (15,743) (8,338 cases), and Camp 15 (13,849 cases). The following camps had the highest case load: Kutupalong RC (33,979 cases), Camp 20 Extension (18,259 cases), Camp 13 (8,338 cases). Therefore, spatial patterns suggest a clustered pattern of distribution which requires a geographical intervention.

**Figure 1.**
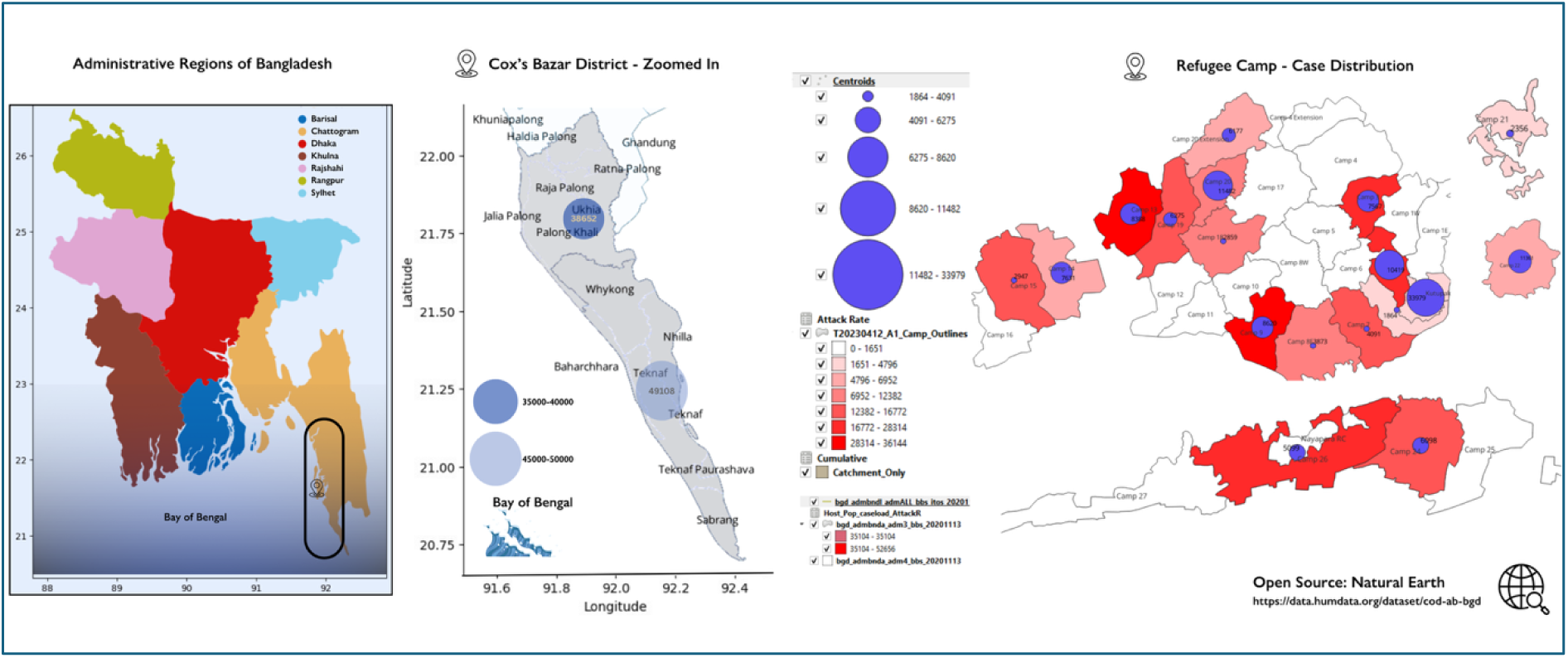
Geo-spatial distribution of scabies cases and attack rates across catchment camps in Cox’s Bazar, Bangladesh (2021–2024).

Table 2 shows that the annual attack rate increased from 538.88 per 10,000 population to a peak of 2283.33 in 2023, which declined to 664.56 in 2024 following the MDA intervention.

**Table 2:** Annual disease burden of scabies and attack rate.

| Year | # of cases | Catchment population | Attack rate (per 10,000) |
| --- | --- | --- | --- |
| 2021 | 32,046 | 594,676 | 538.88 |
| 2022 | 140,590 | 632,634 | 2222.29 |
| 2023 | 150,470 | 658,993 | 2283.33 |
| 2024 | 61,746 | 691,857 | 892.46 |

Figure 2 illustrates the monthly proportional morbidity of scabies in comparison to other reported morbidities from health facilities. The average monthly proportional morbidity of scabies reached a peak of 11.7% in 2023, up from an average of 3.4% in 2021. The average morbidity of scabies again dropped to 5% in 2024 following the MDA campaign.

**Figure 2.**
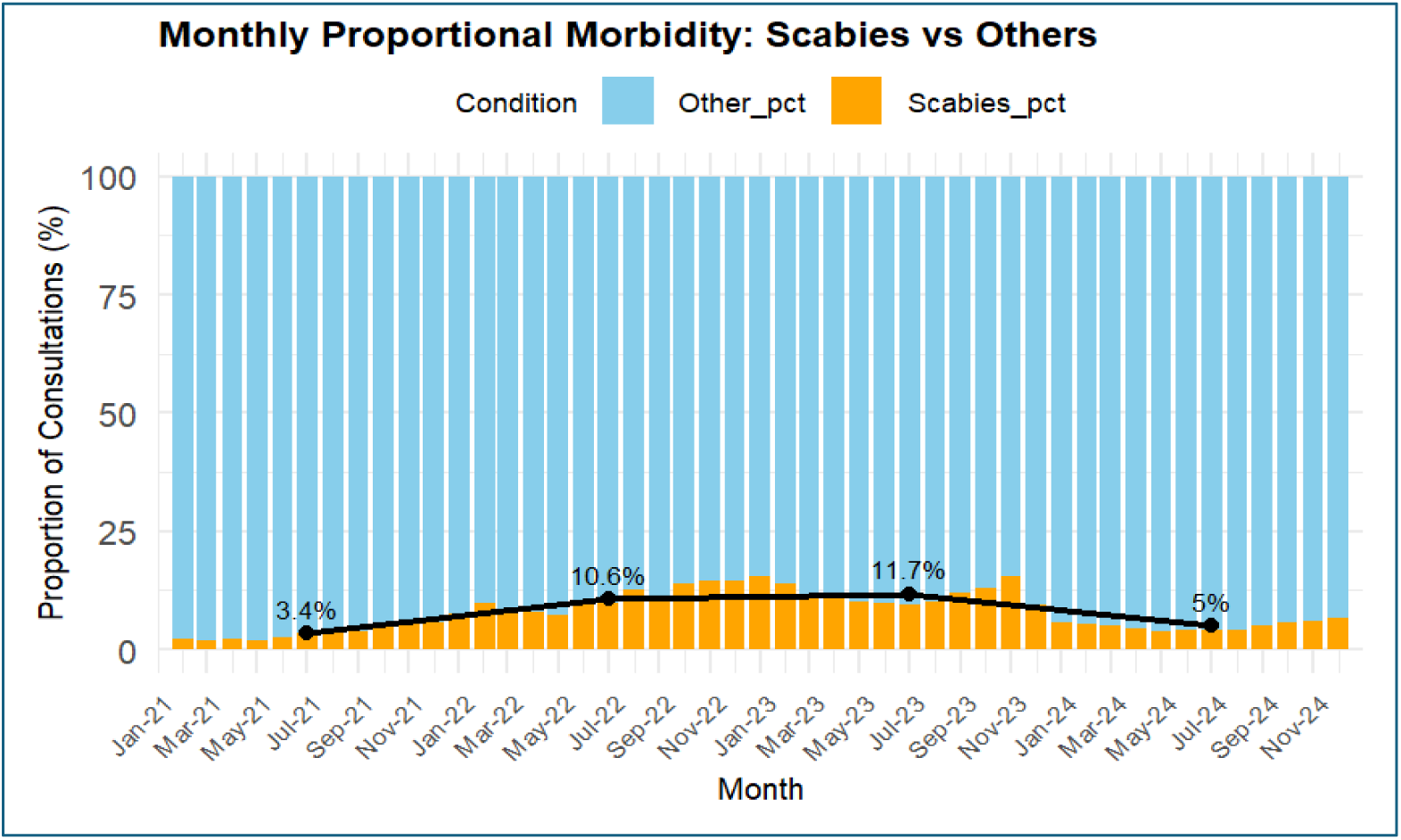
Monthly proportional morbidity (with average morbidity each year): Scabies vs Other Conditions treated at health facilities

Overall, 5.80% of scabies cases were associated with secondary bacterial infections (Table 3). A multivariable regression analysis suggests that male patients had a significantly higher proportion of secondary bacterial infections compared to female patients. The adjusted odds ratio (AOR: **1.00** for males vs **0.86** for females, *p* < 0.001) indicates that females had **14% lower odds**. Children under 5 years of age had a higher proportion of secondary bacterial infections compared to other age groups (7.10% vs 5.06%). Individuals aged **more than 5 years** had significantly lower odds of infected scabies compared to those aged **below 5 years** (Adjusted OR: **0.68**, *p* < 0.001).

**Table 3:** Distribution of scabies by complication (uninfected vs infected scabies) with demographic risk factors.

| Variable | Category | Scabies Count | Infected Scabies Count | Scabies (%) | Infected Scabies (%) | OR | Adjusted OR | $p$ value |
| --- | --- | --- | --- | --- | --- | --- | --- | --- |
| Overall |  | 362532 | 22320 | 94.20% | 5.80% |  |  |  |
| Sex | Male | 167878 | 11039 | 93.83% | 6.17% | 1.00 (ref) | 1.00 (ref) | <0.001 |
|  | Female | 194654 | 11281 | 94.52% | 5.48% | 0.86 | 0.86 |  |
| Age Group (0–5 vs >5) | 0–5 years | 129825 | 9922 | 92.90% | 7.10% | 1.00 (ref) | 1.00 (ref) | <0.001 |
|  | >5 years | 232707 | 12398 | 94.94% | 5.06% | 0.68 | 0.68 |  |

An interrupted time series (ITS) analysis was conducted to evaluate the impact of MDA campaign on the trend of scabies (Figure 3). A significant upward trend was observed from January 2021 until the Mass Drug Administration initiative in December 2023, with an average weekly increase of 23.3 cases (*p* < 0.001). Following the MDA, there was an immediate sharp decline of 1,885 scabies cases per week (*p* < 0.001). The decline sustained for 6 months, with a decline rate of 69.98 per week (*p* < 0.001).

**Figure 3.**
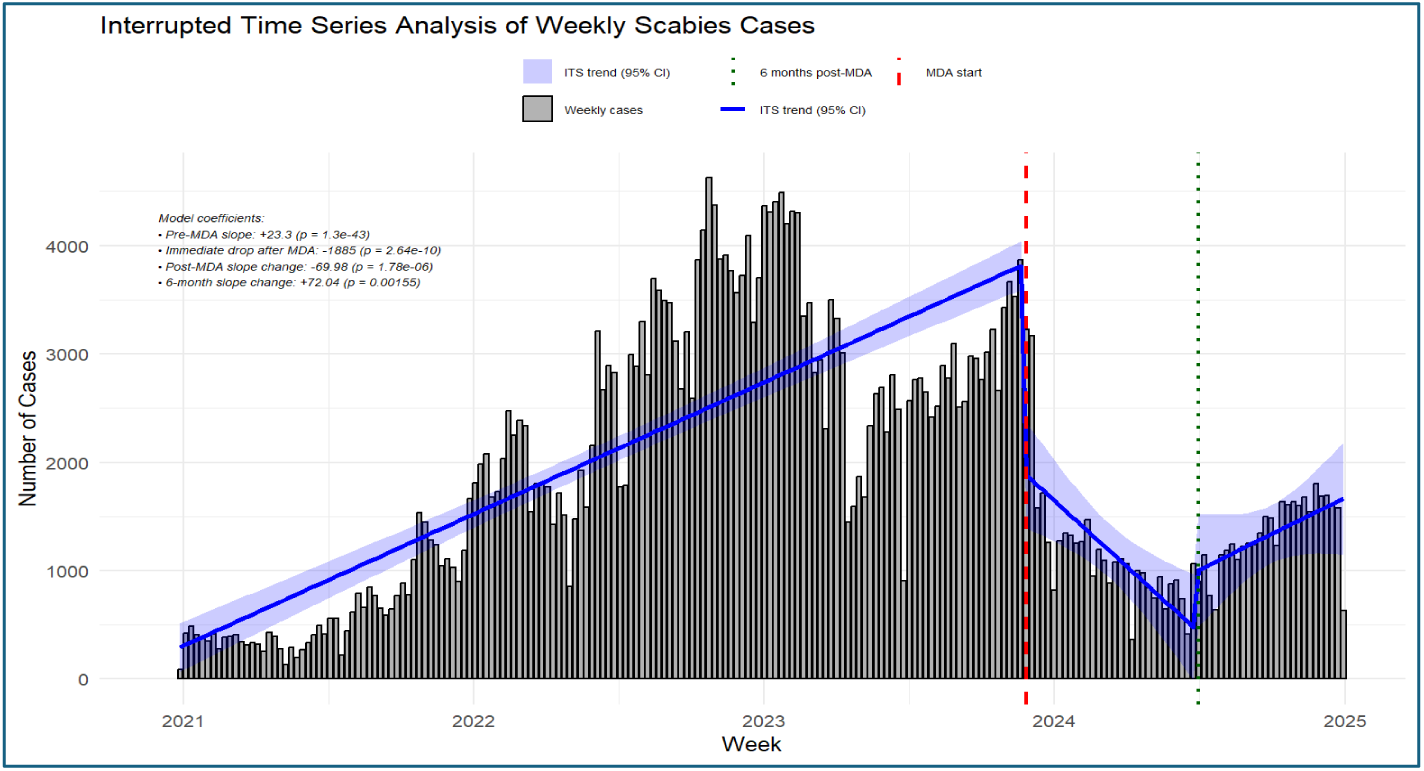
Interrupted Time Series Analysis of Weekly Scabies Trend with pre and post-MDA comparison

To assess the sustainability of the MDA’s impact, the model also included a 6-month post-MDA phase, which revealed a statistically significant upward trend in scabies cases, with an increase of 72.04 cases per week (p = 0.002).

## DISCUSSION

The study found a very high burden of scabies among the Rohingya refugees and adjacent host communities in Cox’s Bazar. It was found that the average annual attack rate of scabies was 1390.65 per 10,000 population, equivalent to 13.91%. Household-level surveys in the same setting found the prevalence to range from 39% to as high as 66<u>%</u> (24). Although the hospital-based attack rate found in the study appears to be lower than that reported in household-level surveys in the same setting, this discrepancy is justifiable because hospital-based data did not capture household contacts/cases not consulted at the health facility level. Scabies, as one of the most prevalent diseases, was also reported in other refugee settings worldwide, including refugee camps in Greece, where 13,118 cases were reported (9) over 4 years, and 2,317 scabies cases from a Turkish hospital (25). However, this study reports one of the highest recorded numbers of scabies cases in a single setting.

The study found the highest burden of cases among children and adolescents, with the highest attack rate among children under 5 years. This finding is consistent with a recent study conducted by MSF at its two hospitals in the same setting (26). A higher susceptibility of children and adolescents to scabies infestation has also been reported in other studies (8,17,20). This may be due to that relatively thinner skin of children makes them more vulnerable to scabies mite penetration, there is an increased likelihood of close skin-to-skin contact among children during play or interactions with household members (17). Such high burden of scabies among children may make them vulnerable to the life-threatening complication of secondary bacterial infections, including acute glomerulonephritis, if untreated (5,6,17,27). The study also found a higher proportion and attack rate of scabies among women in comparison to men, which also aligns with the findings of the studies in similar settings (28). This could be attributed to the fact that women, who often accompany their children to the hospital, are overrepresented in the data.

Scabies accounts for a high proportion of morbidity in IOM supported health facilities in the refugee camps, as high as a monthly average of 11.7%. This proportional burden of scabies in this setting is significantly higher than in similar settings, such as the refugee camp reported in Greece (9). At the same time, scabies can be easily treated with topical permethrin and oral ivermectin; however, such an incredibly high disease burden results in substantial costs in the health sector, placing strain on the limited health resources in the refugee setting. In these contexts, women’s roles as primary caregivers may lead to increased physical contact with children, contributing to higher exposure and transmission risk. Given the recent reductions in donor funding, and the high disease burden caused by scabies in this protracted humanitarian response, a cost-effectiveness analysis of the various treatment regimens is highly recommended to identify low-cost and high-impact treatment approach that is sustainable in this context.

As identified in several studies in the study setting as well as similar refugee contexts worldwide, overcrowded living arrangements, poor hygiene, and lack of water and sanitation facilities are mostly responsible for the high disease burden of scabies in the refugee settings (8,10,17). When traditional case management and hygienic measures fail to prevent and control scabies outbreaks, Mass Drug Administration (MDA) is considered one of the crucial public health interventions in such settings to prevent and control scabies outbreaks among large populations. MDA is a promising approach in outbreak settings for over traditional case-by-case management, because it treats the entire community simultaneously, thereby interrupting the parasite’s transmission cycle (22,29,30).

As the prevalence rate of scabies in the refugee camps surpassed the crucial 10% WHO threshold necessitating MDA, an MDA campaign was conducted from November 29, 2023, to February 01, 2024 (22). The effectiveness of MDA in scabies outbreaks has been reported in different settings, including Australia, the Solomon Islands, Fiji, and the Netherlands (17,29,30). This study also found MDA was effective in reducing annual attack rate and proportional morbidity, with a significant drop in weekly prevalence. Our model also demonstrated that the impact of MDA was sustained in this setting for up to six months, after which the trend began to shift significantly upwards. This implies that without improving living conditions, water, sanitation, and hygiene, MDA alone cannot sustain its impact in the long run. A recent study of IOM, investigated by the same author, revealed the deterioration of water, sanitation, and hygiene in the refugee camps, giving rise to an outbreak of infectious diseases (31). Therefore, urgent attention should be paid to improve the shelter, water, sanitation, and hygiene situation to sustain the effectiveness of mass drug administration, and thereby, prevent the further transmission of scabies. The success of MDA may also be partially attributed to Social and Behaviour Change (SBC) messaging, which played a key role in ensuring treatment compliance and promoting hygiene practices. Sustaining these efforts post-MDA is essential to avoid reinfestation.

Additionally, new refugee influxes during or after MDA implementation could have introduced fresh sources of infestation, contributing to the resurgence observed after six months. Another factor that could contribute to the resurgence of scabies is that the MDA campaign covered only refugees, but not the adjacent host community. As revealed in the study, the attack rate was even higher in the surrounding host community than in the refugee community. The possibility of a parallel surge of scabies with possible transmission between camp and host community was also evidenced in other settings (17,32). Therefore, public health measures should be targeted at both refugees and host communities to halt the transmission of the disease.

## LIMITATIONS OF THE STUDY

The study only included data from IOM-supported health facilities in the refugee camps. Although, as the denominator, we have only taken into account the catchment population of these facilities, the prevalence and attack rate in this study could be underrepresented, as the study could not capture data from facilities run by other partners in the same catchment. While IOM facilities are strategically located throughout the refugee camps, our study only covered areas within their catchment zones and may not be representative of all refugee camps or the host community. Additionally, due to the retrospective nature of the study, we were unable to capture data on some relevant variables, such as behavioral patterns, household contacts, and cases. Despite some limitations, the study provided valuable insights into the epidemiology and magnitude of scabies, as well as the impact of MDA on the outbreak. The study did not attempt to predict the optimal frequency of future MDA rounds, which is critical given the observed resurgence. Further modeling studies are needed to determine appropriate MDA intervals under current camp conditions

## CONCLUSION

The study revealed an extremely high burden of scabies among the Rohingya refugees and the adjacent host community, requiring urgent public health attention. The study reveals the higher vulnerability of children and women, which will necessitate targeted interventions. Furthermore, the study found Mass Drug Administration to be an effective approach for a rapid and substantial reduction in the disease burden. However, the resurgence of the upward trend in scabies indicates that the impact of MDA cannot be sustained if the underlying factors of scabies outbreaks, including overcrowding, water, sanitation, and hygiene, are not addressed. Therefore, we urge policymakers and humanitarian actors to adopt a holistic and coordinated approach to halt the transmission of this disease.

## FUNDING

This study received no specific grant from any funding agency in the public, commercial or not-for-profit sectors.

## COMPETING INTEREST

The authors declare no competing interests. This study received no external funding. The authors used generative AI tools only for language editing and improving clarity during manuscript preparation. All scientific content, data analysis, and conclusions were developed solely by the authors.

## DATA AVAILABILITY

All relevant aggregated data and analysis scripts for this article are available from Zenodo at DOI: https://doi.org/10.5281/zenodo.17687717. The dataset includes anonymized, aggregated scabies case counts and climate variables (temperature, rainfall, humidity) for 2021–2023. Related datasets from the same scabies research series are available at DOIs: https://doi.org/10.5281/zenodo.17697474 and https://doi.org/10.5281/zenodo.17648621.

## ACKNOWLEDGMENTS

CEH conceptualized and designed the study, provided overall methodological guidance, and led the drafting and critical revision of the manuscript. MAH assisted in refining the study design and drafting the manuscript. PPD, MMR, MA, HT, JTP supported field coordination, MAH, PPD, MMR, JPT assisted supporting the data collection processes. MAH, and AAM supported with climatic data triangulation. JCO, ERS, DO, ATMRHB, UMP and JCO contributed to the critical review and refinement of the manuscript. ATMRHB, ERS, UMP, DO and JCO provided strategic oversight and ensured alignment of the study within broader programmatic and policy frameworks. All authors reviewed and approved the final version of the manuscript. CEH is the corresponding author and JCO is the guarantor of the study.

## Notes

### Competing Interest Statement

The authors have declared no competing interest.

### Clinical Trial

N/A ? This study is a retrospective observational analysis and does not involve a clinical trial or any prospective interventional research requiring trial registration.

### Funding Statement

The author(s) received no specific funding for this work.

### Author Declarations

Ethical approval for this retrospective observational study was obtained from the Ethical Review Board of Cox's Bazar Medical College Hospital (Approval Number: CoxMC/2023/017). The analysis used deidentified and anonymized programmatic outbreak surveillance data collected during routine service delivery at IOM-supported health facilities. No identifiable personal information was used, and informed consent was not required due to the retrospective and aggregated nature of the dataset.

## REFERENCES

1. World Health Organization. Neglected tropical diseases – Global [Internet]. [cited 2025 July 26]. Available from: https://www.who.int/health-topics/neglected-tropical-diseases

2. Scabies [Internet]. [cited 2025 July 26]. Available from: https://www.who.int/news-room/fact-sheets/detail/scabies

3. Murray RL, Crane JS. Scabies [Internet]. In: StatPearls [Internet]. Treasure Island (FL): StatPearls Publishing; 2023 [Internet]. [cited 2025 July 26]. Available from: https://pubmed.ncbi.nlm.nih.gov/31335026/

4. IACS [Internet]. [cited 2025 July 26]. International Alliance for the Control of Scabies (IACS). About Scabies. Available from: https://controlscabies.org/about-iacs/about-scabies/

5. Thornley S, Marshall R, Jarrett P, Sundborn G, Reynolds E, Schofield G. Scabies is strongly associated with acute rheumatic fever in a cohort study of Auckland children. J Paediatr Child Health. 2018 June;54(6):625–32.

6. Lynar S, Currie BJ, Baird R. Scabies and mortality. Lancet Infect Dis. 2017 Dec;17(12):1234.

7. Turan Ç, Metin N, Utlu Z. Epidemiological Evaluation of Scabies Cases Encountered in the Last Three Years as a Tertiary Health Center. Turk J Parasitol. 2020 June 1;44(2):77–82.

8. Karimkhani C, Colombara DV, Drucker AM, Norton SA, Hay R, Engelman D, et al. The global burden of scabies: a cross-sectional analysis from the Global Burden of Disease Study 2015. Lancet Infect Dis. 2017 Dec;17(12):1247–54.

9. Louka C, Logothetis E, Engelman D, Samiotaki-Logotheti E, Pournaras S, Stienstra Y. Scabies epidemiology in health care centers for refugees and asylum seekers in Greece. Hay R, editor. PLoS Negl Trop Dis. 2022 June 22;16(6):e0010153.

10. Prevalence of Scabies in Duhok Province and Duhok Refugees’ Camps-Kurdistan Region of Iraq. Int J Sci Technol Res [Internet]. 2020 Oct [cited 2025 July 26]; Available from: https://iiste.org/Journals/index.php/JSTR/article/view/54333

11. United Nations High Commissioner for Refugees (UNHCR). Country – Bangladesh [Internet]. [cited 2025 July 26]. Available from: https://data.unhcr.org/en/country/bgd

12. Polonsky JA, Ivey M, Mazhar MdKA, Rahman Z, Le Polain De Waroux O, Karo B, et al. Epidemiological, clinical, and public health response characteristics of a large outbreak of diphtheria among the Rohingya population in Cox’s Bazar, Bangladesh, 2017 to 2019: A retrospective study. Spiegel P, editor. PLOS Med. 2021 Apr 1;18(4):e1003587.

13. Halder CE, Hasan MA, Mohamud YM, Nyawara M, Okello JC, Mizan MN, et al. Understanding the challenges and gaps in community engagement interventions for COVID-19 prevention strategies in Rohingya refugees: a qualitative study with frontline workers and community representatives. Front Public Health [Internet]. 2023 Aug 3 [cited 2025 July 26];11. Available from: https://www.frontiersin.org/articles/10.3389/fpubh.2023.1169050/full

14. Chin T, Buckee CO, Mahmud AS. Quantifying the success of measles vaccination campaigns in the Rohingya refugee camps. Epidemics. 2020 Mar;30:100385.

15. Halder CE, Hasan MA, Mohamud YM, Nyawara M, Okello JC, Mizan MN, et al. COVID-19 preventive measures in Rohingya refugee camps: An assessment of knowledge, attitude and practice. Dubik SD, editor. PLOS ONE. 2024 Jan 24;19(1):e0282558.

16. Islam MT, Khan AI, Khan ZH, Tanvir NA, Ahmmed F, Afrad MMH, et al. Acute Watery Diarrhea Surveillance During the Rohingya Crisis 2017–2019 in Cox’s Bazar, Bangladesh. J Infect Dis. 2021 Dec 20;224(Supplement_7):S717–24.

17. Engelman D, Fuller LC, Solomon AW, McCarthy JS, Hay RJ, Lammie PJ, et al. Opportunities for Integrated Control of Neglected Tropical Diseases That Affect the Skin. Trends Parasitol. 2016 Nov;32(11):843–54.

18. Campbell M, Van Der Linden N, Gardner K, Dickinson H, Agostino J, Dowden M, et al. Health care cost of crusted scabies in Aboriginal communities in the Northern Territory, Australia. Coffeng LE, editor. PLoS Negl Trop Dis. 2022 Mar 28;16(3):e0010288.

19. Mass Drug Administration (MDAs): an opportunity for community engagement and social behaviour change [Internet]. [cited 2025 July 26]. Available from: https://www.who.int/publications/m/item/mass-drug-administration-(mdas)--an-opportunity-for-community-engagement-and-social-behaviour-change

20. Lake SJ, Kaldor JM, Hardy M, Engelman D, Steer AC, Romani L. Mass Drug Administration for the Control of Scabies: A Systematic Review and Meta-analysis. Clin Infect Dis. 2022 Sept 29;75(6):959–67.

21. Médecins Sans Frontières (MSF). Clinical guidelines: Diagnosis and treatment manual. 2016 ed. Geneva: MSF; 2016.

22. WHO implements large-scale Ivermectin-based MDA for one million Rohingya refugees [Internet]. [cited 2025 July 26]. Available from: https://www.who.int/bangladesh/news/detail/29-01-2024-who-implements-large-scale-ivermectin-based-mda-for-one-million-rohingya-refugees

23. World Health Organization. Scabies [Internet]. [cited 2025 July 26]. Available from: https://www.who.int/news-room/fact-sheets/detail/scabies

24. Rahman MdS, Hasan ABMN, Jahan I, Sharif AB. Prevalence of scabies and its associated environmental risk factors among the Forcibly Displaced Myanmar Nationals living in the Cox’s Bazar district of Bangladesh. J Migr Health. 2024;9:100220.

25. Coşkun Ö, Aynure Ö, Engin Ş, Güreser Ayşe Semra, Agah TM, Taylan ÖA. Retrospective analysis of scabies cases admitted to a Turkish hospital according to the citizenship status of the patients. 2022 Oct 26 [cited 2025 July 26]; Available from: https://zenodo.org/record/7250314

26. Alhaffar BA, Islam S, Hoq MI, Das A, Shibloo SM, Hasan M, et al. High caseload of Scabies amongst Rohingya refugees in Cox’s Bazar, Bangladesh: A retrospective analysis of the epidemiological and clinical characteristics of cases, July 2022 to November 2023. Henao-Martínez, AF, editor. PLOS Glob Public Health. 2025 Apr 9;5(4):e0003981.

27. Chisavu F, Gafencu M, Steflea RM, Vaduva A, Izvernariu F, Stroescu RF. Acute Glomerulonephritis Following Systemic Scabies in Two Brothers. Children. 2024 Aug 14;11(8):981.

28. Schneider S, Wu J, Tizek L, Ziehfreund S, Zink A. Prevalence of scabies worldwide—An updated systematic literature review in 2022. J Eur Acad Dermatol Venereol. 2023 Sept;37(9):1749–57.

29. Van Deursen B, Hooiveld M, Marks S, Snijdewind I, Van Den Kerkhof H, Wintermans B, et al. Increasing incidence of reported scabies infestations in the Netherlands, 2011–2021. Mossong J, editor. PLOS ONE. 2022 June 24;17(6):e0268865.

30. Romani L, Whitfeld MJ, Koroivueta J, Kama M, Wand H, Tikoduadua L, et al. Mass Drug Administration for Scabies — 2 Years of Follow-up. N Engl J Med. 2019 July 11;381(2):186–7.

31. International Organization for Migration (IOM). Rise of Hepatitis A in Rohingya Refugee Camps: A qualitative investigation of the early surge. Available from: https://reliefweb.int/report/bangladesh/rise-hepatitis-rohingya-refugee-camps-qualitative-investigation-early-surge.

32. Romani L, Steer AC, Whitfeld MJ, Kaldor JM. Prevalence of scabies and impetigo worldwide: a systematic review. Lancet Infect Dis. 2015 Aug;15(8):960–7.

